# Moving from GWAS signals to rare functional variation in inflammatory bowel disease through application of GenePy2 as a potential DNA biomarker

**DOI:** 10.1101/2024.04.19.24306093

**Authors:** Guo Cheng, James J Ashton, Andrew Collins, R Mark Beattie, Sarah Ennis

## Abstract

**Objectives:** We adopt a weighted variant burden score GenePy2.0 for the UK Biobank phase 2 cohort of inflammatory bowel disease (IBD), to explore potential genomic biomarkers underpinning IBD’s known associations.

**Design:** Nucleating from IBD GWAS signals, we identified 794 GWAS loci, including target genes/LD-blocks (LDBs) based on linkage-disequilibrium (LD) and functional mapping. We calculated GenePy2.0–a burden score of target regions integrating variants with CADD_Phred_≥15 weighted by deleteriousness and zygosity. Collating with other burden-based test, GenePy-based Mann-Whitney-U tests on cases/controls with varying extreme scores were used. Significance-levels and effect sizes were used for tuning the optimal GenePy thresholds for discriminating patients from controls. Individual’s binarized GenePy status (above or below threshold) of candidate regions, was subject to itemset association test via the sparse Apriori algorithm.

**Results:** A tailored IBD cohort was curated (n_Crohn’s_Disease(CD)_=891, n_Ulcerative_Colitis(UC)_=1409, n_Controls_=60118). Analysing 885 unified target regions (794 GWAS loci and 104 monogenic genes with 13 overlaps), the GenePy approach detected statistical significance (permutation *p*<5.65×10^-5^) in 35 regions of CD and 25 of UC targets exerting risk and protective effects on the disease. Large effect sizes were observed, *e.g. CYLD-AS1 (*Mann-Whitney-□=0.89[CI:0.78-0.96]) in CD/controls with the top 1% highest scores of the gene. Itemset association learning further highlighted an intriguing signal whereby GenePy status of *IL23R* and *NOD2* were mutually exclusive in CD but always co-occurring in controls.

**Conclusion:** GenePy score per IBD patient detected ‘deleterious’ variation of large effect underpinning known IBD associations and proved itself a promising tool for genomic biomarker discovery.

**What is already known on this topic:** Inflammatory bowel disease (IBD) is a genetically heterogeneous disease with both common polygenic, and rare monogenic, presentations. Previous studies have identified known genetic variants associated with disease.

**What this study adds:** A genomic biomarker tool, tailored for large cohort, GenePy2.0 is developed. It’s rank-based test is more powerful than mutation-burden based test in validating known associations and finding new associations of IBD. We identified large risk and protective effects of ‘pathogenic genes/loci’ in IBD, including expanding previous associations to wider genomic regions.

**How this study might affect research, practice or policy:** GenePy2.0 facilitates analysis of diseases with genetic heterogeneity and facilitates personalised genomic analysis on patients. The revealed genetic landscape of IBD captures both risk and protective effects of rare ‘pathogenic’ variants, alongside more common variation. This, could provide a fresh angle for future targeted therapies in specific groups of patients.

## INTRODUCTION

Inflammatory bowel disease (IBD) is a chronic, highly heterogenous, inflammatory condition resulting from an aberrant immune response to environmental triggers, in genetically susceptible individuals[1]. The disease is commonly classified into two subtypes: Crohn’s disease (CD) and ulcerative colitis (UC) based on clinical findings, yet the clinical phenotype is much more varied and hinders effective disease treatment.

Genome-wide association studies (GWASs) of common variants (minor allele frequency (MAF)>3∼5% in the general population) on IBD, driven by large cohorts including the UK Biobank[2], have identified over 300 IBD-associated loci, which shed light both on the IBD genetic landscape and implicate pathways[1]. However, with modest effect sizes, common variants altogether explain only a minor fraction of the observed IBD heritability, and most of the GWAS variants are not causal variants but rather proxies of the causative variations based on linkage disequilibrium (LD)[3]. This limits the application for clinical translation of GWAS.

Rare variants (RVs) with major effect sizes represent a key genomic driver that have known associations with IBD[4, 5]. Although methodologically challenging[4], a growing number of RVs have been statistically identified in complex IBD and functionally validated for monogenic IBD, with the latter manifesting as a rare Mendelian subtype of IBD with familial clustering of occurrence, often with specific additional features, such as immunodeficiency[6, 7]. Association of RVs often overlap with common variants [8]. In complex IBD, evidence suggests these variants may be associated with disease through a 2^nd^ hit mechanism by RV in the pathogenesis pathway/gene in addition to a GWAS association, or via synthetic association of RVs underpinning the GWAS signal in the LD region, while both mechanisms may hold the key to decipher the RVs’ role in disease[3, 9–11]. The means that both targeted genes of the GWAS association and the region delineated by the LD block (LDB) that encompasses the association signal can host the disease causal RVs. LDBs can vary among different ethnic groups, whilst targeted genes either under influence of expression Quantitative Trait Loci (eQTL) or in physical adjacency of the variant cannot always be clearly defined due to pleiotropic effects and epigenetic modifications[12–15].

Within a targeted genomic region, burden-based test of selected RVs, *e.g*. missense variants, or loss-of-function variants, is the norm to check for the case versus control associations, with well-established sequence kernel association test (SKAT/SKAT-O) for example. However, refined pathogenicity weighting of the variants in the burden tests can be essential to elucidate the role of RVs in GWAS loci and in disease. Taking the ‘mendelian-complex’ genes, the causal genes that overlap between complex IBD and monogenic IBD, for instance, while damaging mutations of these genes cause a severe phenotypic presentation as monogenic IBD, the variant of modest effect can predispose risk to a milder polygenic form of the phenotype as identified from GWASs[6]. The pathogenicity variance of the variants in the same gene is the cause of the vastly different phenotypic presentation in this case. Methods based on this, represented by GenePy score integrating mutation load, allele frequency and pathogenicity score of individual variants, has been successfully applied in both clinical genetics and machine learning models based on small cohorts of data[16, 17].

In this study we developed GenePy2 to adapt with large cohort of rare variants data and tested it as a prototype of a DNA biomarker for IBD. This was followed by investigations on disease association and personalized examination on patients’ genetic landscape of disease. Analyses were carried out on the UK Biobank cohort and tested on GWAS association regions of IBD.

## MATERIALS AND METHODS

### The UK BioBank IBD cohort

The analysis is based on the UK Biobank phase 2 dataset (project 72911), encompassing exome sequencing and detailed phenotype information from approximately 200,000 participants, which was publicly released in October 2020[18, 19]. Participants who have withdrawn were excluded from the analysis. The exomes were captured using the IDT xGen Exome research panel V1.0, designed to target 39 Mbps of the human genome. To ensure data quality, additional quality control (QC) metrics were applied to the project-VCF (pVCF). A detailed workflow of this process, along with a list of immune-related diseases curated by the clinic and informatics team, is presented in Figure 1a and supplementary methods with Table S1.

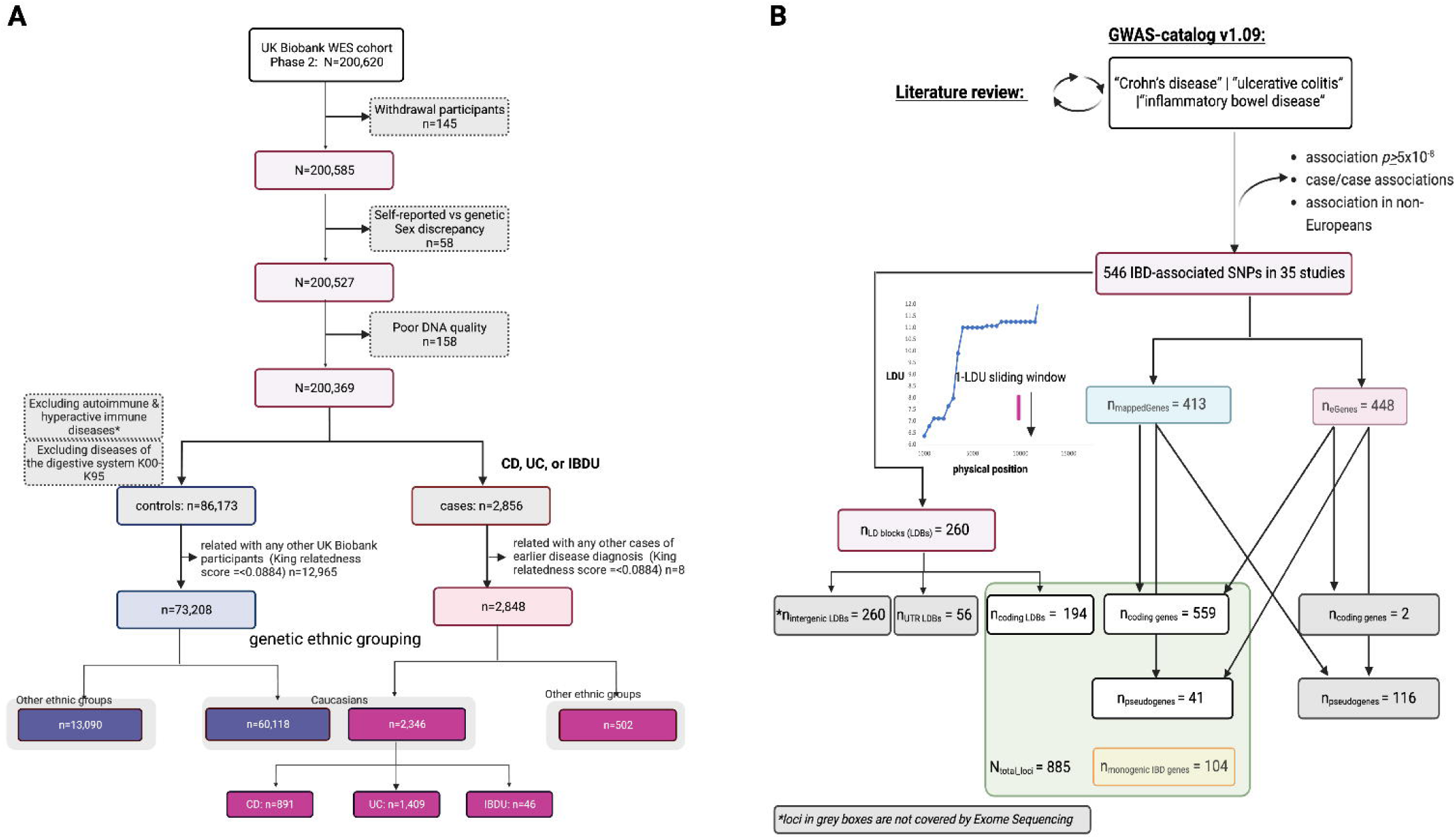

Patients or the public WERE NOT involved in the design, or conduct, or reporting, or dissemination plans of our research.

### Curation of the IBD-associated genomic variants

UC, CD, or IBD-associated Single-nucleotide polymorphisms (SNPs) with maximum association p-value of 5×10^-8^ were retrieved from the GWAS Catalogue v1.09 [20]. Through a literature review conducted in June 2023, we refined the dataset by excluding associations derived from case-case studies, associations related to disease subtypes other than UC and CD, as well as those identified in non-European populations. The PubMed search query utilized for literature review was: ((((((“Crohn’s disease”) OR “inflammatory bowel disease”) OR “Ulcerative Colitis”)) AND ((“genome-wide association“[Title/Abstract]) OR “genome-wide association“[Title/Abstract]))) AND (“1000“[Date - Create] : “2023/06/07“[Date - Create]).

For each association SNP, we first examined its physically mapped genes (mappedGenes), using the same approach of GWAS Catalogue[20]. We identified cis-regulated genes (eGenes) using data from the recent GTEx database V8 (https://www.gtexportal.org/), by extracting those associated-SNPs that function as expression Quantitative Trait Loci (eQTLs) in tissues including transverse colon, sigmoid colon, small intestine&terminal ilium, EBV-transformed lymphocytes, fibroblasts, and whole blood [21].

To delineate linkage disequilibrium (LD) blocks (LDBs) associated with IBD, we projected the locus of each association SNP onto the LD unit map of the European population. Employing a sliding window of 1 LD unit (LDU) in size, whereby loci within a 1-LDU distance are grouped into one LDB [14, 15]. In cases where direct interpolation of a locus was not feasible, we utilized the position of the most adjacent marker for this purpose. Such LDBs and targeted genes were identified as GWAS loci in this study.

Monogenic IBD genes retrieved from literature are also included in the analysis. The curation process of all the candidate loci is illustrated in Figure 1B.

### Per-locus GenePy score for the IBD cohort

The GenePy v2.0 was developed to cope with large cohort data by addressing issues: 1) incorporation of multi-allelic considerations into the score (maximum-n_alternative_allele_=10); 2) enabling the calculation of scores for various genomic regions, such as LDBs; 3) computational cost reduction with optional processing using GPU; 4) optional selection of variants that are pathogenic or likely pathogenic. The score was built on assessing the pathogenicity potential of each variant allele besides the variant load of a genomic region, integrating information from the Combined Annotation–Dependent Depletion(CADD V1.6) score and population allele frequency as observed in the 200K participants [22]. Genepy2 score was calculated for each candidate gene or LDB based on likely-pathogenic variants of CADD_phred_score_ ≥15 for every individual within the cohort.

Details of calculation were described in supplementary methods. The GenePy2.0 pipeline is open source and can be accessed at https://github.com/UoS-HGIG/GenePy-2

### LDB/gene-based mutation test

GenePy2 score-based Mann-Whitney U test was conducted with other burden and threshold-based tests (supplementary methods). We considered the genetic heterogeneity of IBD, with the most commonly associated gene, *NOD2* for example, estimated to account for 7.5% of Crohn’s disease cases [7, 23] therefore tapered the test from all individuals to those with all non-zero score, top 7.5%, 5%, 2.5% and 1% of highest GenePy scores in cases and controls respectively, to provide a more statistically robust characterisation of the contribution of each gene to disease pathogenesis. This was also followed by a permutation test of 10^5^ times to address confounding effects caused by population stratifications. The effect size of the Mann-Whitney U test was evaluated using the Mann-Whitney parameter, theta

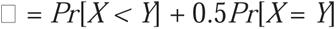

with bootstrap resampling to estimate its confidence interval[24–26].

All associated tests utilized the same sets of variants and identical LDB/gene coordinates, specifically focusing on pathogenic variants with a CADD_Phred_Scor<u>e</u>_<u>></u>15. Mann-Whitney U tests are based on the scikit-learn library of Python 3.7[27].

### Itemset analysis

GenePy status was defined as follows: individuals in the sub-population (*i.e.* those with the top 7.5%, 5%, 2.5%, or 1% highest scores, or all with non-zero scores) whereby maximum effect size is observed in the GenePy-based Mann-Whitney U test, were deemed positive for GenePy for the testing locus, others with lower scores as negative. The binarization process was conducted for UC case/controls and CD case/controls separately. GenePy status of associated loci (Mann-Whitney U test permutation *p*<5.65×10^-5^ addressing multiple testing issue), was analysed by an item association rule mining unsupervised learning approach via the APRORI algorithm[28, 29], as implemented in arules and arulesViz package of R[30, 31]. To reduce the sparsity of the data, individuals without any positive GenePy status were removed before the association mining.

Itemset support (*i.e.* frequency), lift, and confidence were examined in both CD and UC cohorts and for cases and controls separately to understand the pattern of co-occurrence of association loci, exploring the potential epistatic effects of risk and protective variants. The minimum itemset support for the analysis was 0.0001, and minimum confidence was 0.5.

## RESULTS

### LDB and genes in association with IBD

A total of 546 IBD-associated single-nucleotide polymorphisms (SNPs) were identified from 35 association studies (Table S2), corresponding to 718 GWAS genes. This set includes 413 mappedGenes and 448 eGenes, with an overlap of 143 genes, as depicted in Figure 1B and Table S2. Notably, 13 of the 104 monogenic IBD genes (monoGenes) are GWAS genes, i.e. ’Mendelian-complex genes’, exhibiting significant intersection (Fisher’s exact test; protein-coding genes only, *p*=6.72×10^-6^). Functional gene set enrichment analysis revealed similar enrichment of both GWAS genes and monoGenes in immune-related pathway (Table S3; Figure S1), aligning with the anticipated convergence of molecular pathogenic pathways in monogenic IBD and complex IBD.

Another feature of GWAS genes is the enrichment of non-protein coding pseudogenes (n=157), which make up 26.39% of mappedGenes and 12.95% of eGenes. This aligns with overrepresentation of pseudogenes in the applied reference GENCODE V43 [32, 33] and our impartial SNP-gene mapping approach with no preferable selection for protein-coding genes or known IBD genes. Whilst there is growing knowledge of their association with disease and immune regulation [34], the majority of the pseudogenes are not covered by the Exome capture kit (n=116; Figure 1b).

Utilizing the European-based LDU map[14], 546 GWAS SNPs are categorized into 260 LDBs, with 150 consisting of a single association SNP (IBD-association *p*<5×10^-8^), and the remaining defined by ≥ 2 GWAS association SNPs. The LDBs span from 1.00 to 3.20 LDU, or 3,630bp to 3,246,717 bp according to the physical position in size. The largest LDB, LDB78b, is located at 5q31.1 (Table S2), and encompasses 6 GWAS association SNPs, which consist of eQTLs of *MEIKIN*. LDB78a, despite being >1LDU far away from LDB78b, encompasses another IBD-associated eQTL of *MEIKIN*. Such LDBs, by sharing a common gene with the association SNP that they encompass, are defined as clusters of LDBs (n=21). As might be expected, the most significant cluster is the HLA region at 6p21.32-33, comprising 7 LDBs (Table S2). One hundred and ninety-four LDBs are captured by the Exome sequencing assay. These LDBs encompass the complete sequence of 313 GWAS genes, partially overlap with 201 GWAS genes, and have no intersection with the other 204 GWAS genes. LDBs can also extend beyond mappedGenes and eGenes. For instance, LDB187 at 16q12.1, delineated by 5 GWAS SNPs covers *CYLD*, a monoGene but not a GWAS gene, besides *NOD2* and *CYLD-AS1* (Figure S2).

The GWAS genes, LDBs and monoGenes together account for 885 target regions to be tested, as component LDBs within a LDB cluster is tested separately.

### The UK Biobank IBD cohort

Following QC, ethnicity- and phenotype-based filtration retained 891 CD, 1,409 UC cases, and 60,118 controls. Most of the IBD diagnoses were made in patients’ adulthood, whilst 37 CD and 33 UC were diagnosed on or before the patients reached 18 years old. Further demographic and sub-phenotypic features of the UC and CD patients are derived based on the ICD-10 code of diagnosis as shown in Table 1.

**Table 1.** Characteristics of the European UK BioBank cohort for the analysis.

|  | CD (n=891) | UC (n=1409) | Controls (n=60,118) |
| --- | --- | --- | --- |
| **Demographics** |  |  |  |
| Male | 387 (43.43%) | 730 (51.81%) | 32925 (54.77%) |
| Female | 504 (56.57%) | 679 (48.19%) | 27193 (45.23%) |
| Age at latest assessment: median (IQR) (Year) | 59 (51-64) | 61 (54-65) | 58 (50-63) |
| Age at Diagnosis; median (IQR) (Year) | 51 (34-62) | 52 (37-63) | NA |
| **Disease Subtypes** |  |  |  |
|  | Small intestine<br>Large intestine<br>small & large intestine<br>Unspecified | ileocolitis<br>proctitis<br>rectosigmoiditis<br>proctus &<br>rectosigmoiditis<br>unspecified | NA |
|  | 174<br>197<br>47<br>473 | 6<br>192<br>81<br>25<br>1105 | NA |
| **GI complications** |  |  |  |
| Fistula disease | 57 (6.40%) | 30 (2.13%) | NA |
| Stricture disease | 138 (15.49%) | 57 (4.05%) | NA |
| colon cancer | 26 (2.92%) | 43 (3.05%) | NA |
| megacolon disease | 4 (0.45%) | 6 (0.43%) | NA |
| **Comorbidities with other autoimmune diseases** |  |  |  |
| n=1 | 111 (12.46%) | 151 (10.72%) | NA |
| n=2 | 15 (1.68%) | 15 (1.06%) | NA |
| n>=3 | 1 (0.11%) | 5 (0.35%) | NA |

### Pathogenic mutations of GWAS association loci and monogenic IBD genes

All but 10 of the GWAS-derived set of 794 targets host ≥1 variants with CADD_phred_score_≥15 in the cohort, and all the monoGenes were mutated in at least 1 patient. Despite this, pathogenic variants were very sparsely identified in the patients. Approximately half of the testing loci had a non-zero GenePy score in fewer than 5% of patients, as observed on 416 (52.39%) of the GWAS loci and 46 (44.23%) of the monoGenes in CD patients, and similarly on 425 (53.53%) GWAS loci and 48 (46.15%) monoGenes in UC. With more than half of the values being zeros, the GenePy score matrix per locus/individual is a sparse matrix for downstream analysis.

The most mutated genes are the 13 known ‘mendelian-complex’ IBD genes, as 8 (61.53%) are mutated in >5% of both UC and CD, except for *CD40, IL2RA, IL10, STAT3* and *LACC1* that are rarely mutated either UC or CD. Such sparsity of non-zero GenePy scores of the patients corresponds to the genetic heterogeneity of IBD and is the rational for the following GenePy-based association tests on subpopulations with highest scores.

### Association of the candidate regions with disease

Under the monogenic IBD model, two significant associations are observed with CD which exert opposing effects on disease: *NOD2* being risk under the recessive model and *IL23R,* under the dominant and additive inheritance models, both with protective effects. Both genes are known IBD genes with *NOD2* also being a ‘mendelian-complex’ IBD gene. No significant associations were detected with UC from this test (Figure S3).

Burden-based SKAT-O test highlighted the most significantly associated gene of UC, *RIPK2-DT*, a noncoding eGene associated with the IBD-association SNP rs7015630. *RIPK2-DT* plays a role in mitigating inflammation induced by free fatty acids but is less known in IBD compared to its downstream gene *RIPK2* [35, 36]. The *RIPK2-DT* association was not detected in the GenePy-based rank sum tests.

GenePy-based Mann-Whitney U test uncovered 35 loci in significant association with CD and 25 with UC (Figure 2; Figure S4). *HLA-DQA1* and *HLA-DQB2* are the most significantly associated genes with UC and controls of the top 7.5% or GenePy scores, albeit of modest effect sizes (□*_HLA-DQA1_*=0.63, CI [0.59,0.67]; □*_HLA-DQB2_*=0.66, CI [0.63,0.70]), compared to other associated genes, *e.g*. □*_SLC17A1_*=0.81, CI [0.73,0.88], or the monoGene *LIG4*, where □*_LIG4_*=0.82, CI [0.74,0.89]; *NOD2*, together with the co-located LDB187 and *CYLA-AS1* gene (Figure S2), but not *CYLD* the monoGene, are the most significantly associated with CD (Figure 2). Such associations propped up by the rare pathogenic variants (CADD_phred_score_≥15) exert larger effect sizes to disease compared to that identified from the original GWAS, of both protective and risk effects observed, and such effects tend to be bigger when the afftected sub-population is smaller (Figure 2). Notably, although the smallest p value of NOD2 was observed in individuals of the top 7.5% highest scores (*p*=1.41×10^-17^, □=0.80, CI [0.77,0.83], the maximum effect size was observed in those with the top 2.5% highest score *p*=5.13×10^-7^, □=0.81, CI [0.76,0.86])

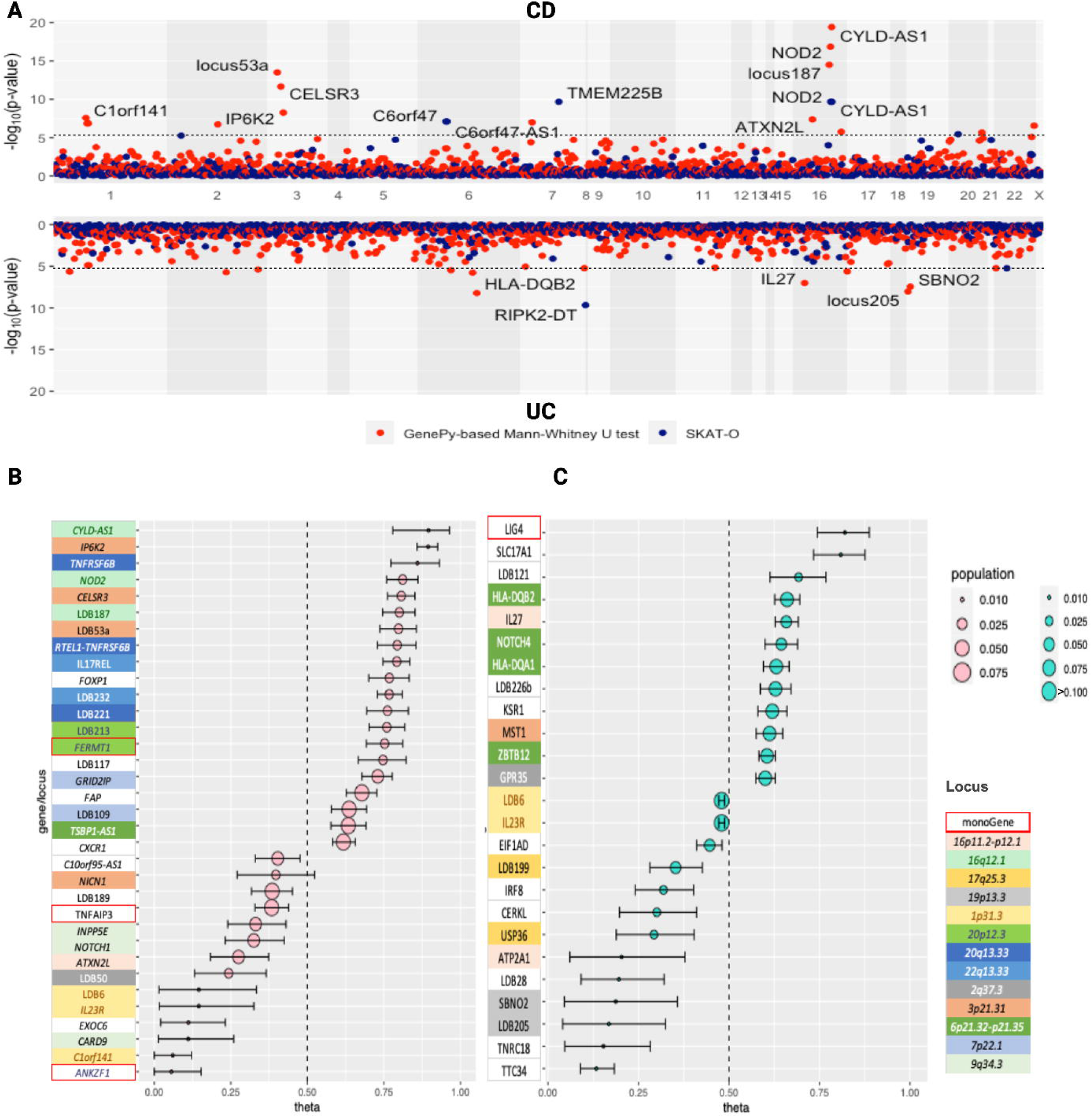

The eGene *NOTCH1* and a mappedGene *CARD9* at locus 9q34.3 are tagged by the association SNPs encompassed by LDB131a/b, and both exhibited significant association with CD, evincing pleiotropic effects at the gene level of a GWAS association locus. In another case, LDB189 which constitutes a proportion of the *PLCG2* gene encompassing the phospholipases domain, is significantly associated with CD with protective effects but the entire *PLCG2* gene is not (Figure 3), in line the GWAS findings[37].

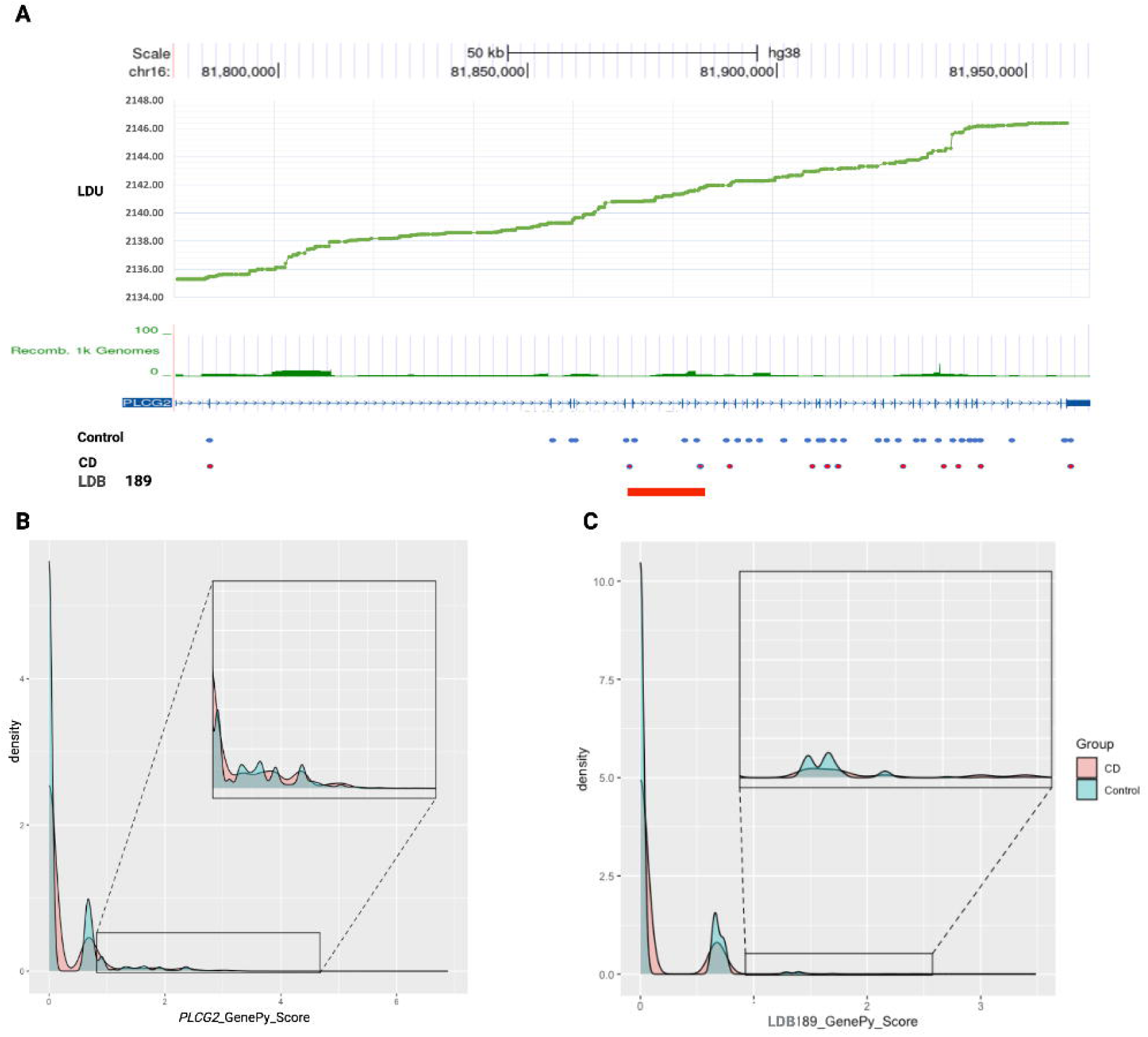

### Set of highly mutated genes in IBD and controls

We tested Rare variant-based associations in both UC and CD, appeared to exert both protective and risk effects, with the potential for some cases of the disease to constitute oligogenic pathogenesis given the large effect sizes. We tested this using an itemset association analysis by the APRIORI algorithm, with patients carrying higher GenePy2 score than the cut-off applied in association tests considered being GenePy positive for a mutant gene or LDB (Table S5-8). The test was conducted on 398 CD patients and 28,017 controls with any positive GenePy status of the 34 CD-associated genes/LDBs, and similarly on 613 UC with 25,748 controls with positive GenePy of the 25 association loci.

GenePy status of LDBs/genes within the same GWAS association region tend to be associated because of the existing intra-region overlaps (Table S2 and Table S5-8). Between GWAS regions, considerable coexistence of ‘positive’ GenePy status of LDB187/*NOD2* and *IR23R*/LDB6 were observed in controls (Figure 4A). This coexistence was completely absent in itemset observation in CD cases, with GenePy(+) status of both the *NOD2/IL23R* regions being mutually exclusive in CD patients (Figure 4A; Table S5-S6). *IL23R* and the genomic region also showed strongest associations with other regions in controls of the UC-associated genes/LDBs (Figure 4B; Table S7-8), indicating its counter-risk effects in both IBD subtypes, albeit the observation in UC can be biased as the sub-population with *IL23R* positive GenePy status constituted 14.08% of the UC cohort (Figure 3C).

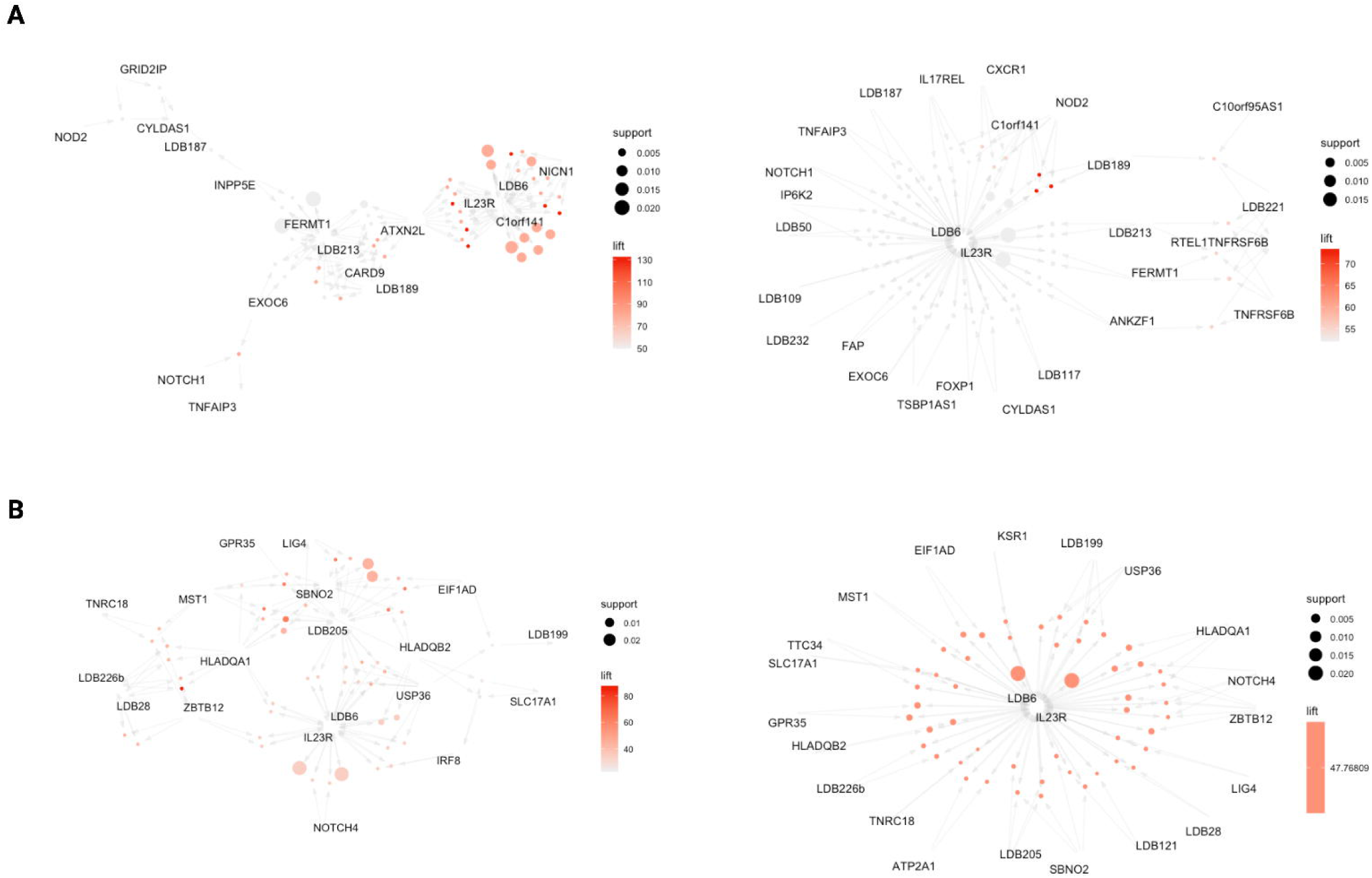

## DISCUSSION

Both single DNA variant and aggregated effects of multiple variants has been utilized for disease risk stratifications[38, 39], but a biomarker from rare and functional genomic variants is missing for complex disease despite their potentially direct causal effect with disease. Filling the gap relies on a large cohort, but big genomic data is enriched with issues of complex variations, e.g. multi-allelic variation, variation of unknown significance, etc. Based on the UK BioBank cohort, we tackled such complexities using an evolved GenePy2.0, and then tested it on known GWAS loci represented by common variants-based associations. A tailored analysis on IBD was performed, and the result demonstrated the significant enrichment of associations represented by GenePy score with both risk and protective effects on disease occurrence, which will change our previous outlook on the IBD genetic architecture. This approach also exemplifies a new approach to tackling the relationship of GWAS CVs and rare variants.

IBD is the archetypal ‘complex’ disease, with genetic heterogeneity leading to distinct underlying aetiology of disease pathogenesis within individuals and governing the role of both triggering and ongoing environmental drivers of disease. In addition to the plethora of GWAS findings which shed light on the genetic pathogenic pathways of the disease, recent analysis of large numbers of patients with WES data has continued to advance knowledge and implicated more rare variation in pathogenesis [4]. Here we build upon the ability to assess rare variation through application of statistical analysis to determine the maximal contribution of each GWAS locus to IBD pathogenesis within the cohort.

We did not limit our view on GWAS to genes, instead followed a naïve approach revisiting the SNP associations with LD mapping in addition to evidence-based physical and eQTL mapping of candidate genes. This introduced pseudogenes and intergenic LDBs which are undesirable targets for the WES-based downstream analysis as many of the candidates are not captured by the sequencing assay, not to mention that many are less studied. However, this has also led to novel discoveries. Our analysis points to variation across the entire *NOD2*-associated LDB, rather than just the gene, as being significantly associated with Crohn’s disease inferring important roles for regulatory regions in addition to established coding variants. Similarly, our analysis pinpoints an association in the *PLCG2* to only part of the gene with the potential to utilise this to better understand the underlying biological process through which variants lead to disease. Pseudogenes *RIPK2-DT* and *CYLD-AS1* also stand out in association tests which indicates novel pathogenic gene pathway of IBD.

The discovery of associations has been significantly promoted by GenePy. By capturing the role of rare variation at an individual level, this technique provides the ability to both determine the relative contributions to IBD pathogenesis of associated genes across a cohort, and to determine, at an individual level, patients presumed to have disease where a specific gene (or set of genes) has a statistical contribution compared to other patients. This opens the possibility of personalising the molecular diagnosis for an individual patient and identifies genomic biomarkers of disease. By taking the subset of individuals with the highest GenePy score, we can tackle the genetic heterogeneity of IBD in a straightforward approach. For instance, rank-based comparison recovered the most significant association of *NOD2* locus for the CD patients with the top 7.5% GenePy scores, concurring with previous findings, although we found that the largest hazard effect of *NOD2* mutation is for the more extreme top 1% of scores.

Not all ‘pathogenic’ variants are causal to IBD, as we found both risk and protective effects in the CD and UC cohort. This is consistent with the evolutionary picture of autoimmune disease[40], and the directionality of genetic variants may be addressed in burden-based association test able to annotate gain-of-function, loss-of-function and dominant negative effects into the GenePy score in the near future. Interestingly, the effect sizes of the GenePy score based tests are much larger than the GWAS findings on index SNPs, providing the possibility for the scoring tool to be applied as a potential biomarker for implicated genomic ()counter each other when occurring to the same individual, as we observed in controls with positive *NOD2* GenePy status being also positive for *IL23R*. Furthermore, identifying this pattern implicates an oligogenic picture of IBD for some patients, with disease aetiology lying between complex and monogenic IBD.

Whilst the UK Biobank cohort provides many advantages, including its large size and rich phenotyping data, the nature of WES data are not ideal for analysis of all GWAS targets as many of the associations lie in noncoding regions, as observed in a large proportion of the LDBs in this study. WGS may provide the opportunity for improvement in both methods and discovery, and application of these methods. Another area of potential weakness in UK Biobank data are the precision of the clinical phenotyping, which impedes the subtype or genotype-phenotype correlation analysis even with GenePy of large effects. In this study we have attempted to identify genomic associations of specific IBD subtypes and are therefore reliant on the accuracy of clinical data to make correct associations. It is also important to recognise that quality control of phenotypes by specific researchers is not possible and we have used the available data to categorise IBD patients, and to identify controls that are reported to have no other autoimmune conditions.

With approved access to the Phase 3 UK Biobank data (project ID140070) and other IBD data, we are looking to replicate the GenePy-based findings on IBD and other diseases, with testing and development of GenePy as a potential DNA biomarker representing rare functional variants for complex diseases.

## Data Availability

All data produced in the present work are contained in the manuscript

## Acknowledgements

This study is funded by AGENDA EPSRC funding on AI health research (EP/Y01720X/1) and was supported by the National Institute for Health Research (NIHR) Southampton Biomedical Research Centre. The views expressed are those of the author(s) and not necessarily those of the NIHR or the Department of Health and Social Care. The authors acknowledge the use of the IRIDIS High Performance Computing Facility, and associated support services at the University of Southampton, in the completion of this work. JJA is funded by an NIHR Advanced fellowship.

## REFERENCE

1. Graham, D.B. and R.J. Xavier, Pathway paradigms revealed from the genetics of inflammatory bowel disease. Nature, 2020. 578(7796): p. 527–539.

2. Jiang, L., et al., A generalized linear mixed model association tool for biobank-scale data. Nat Genet, 2021. 53(11): p. 1616–1621.

3. Uffelmann, E., et al., Genome-wide association studies. Nature Reviews Methods Primers, 2021. 1(1).

4. Sazonovs, A., et al., Large-scale sequencing identifies multiple genes and rare variants associated with Crohn’s disease susceptibility. Nat Genet, 2022. 54(9): p. 1275–1283.

5. Gettler, K., et al., Common and Rare Variant Prediction and Penetrance of IBD in a Large, Multi-ethnic, Health System-based Biobank Cohort. Gastroenterology, 2021. 160(5): p. 1546–1557.

6. Bolton, C., et al., An Integrated Taxonomy for Monogenic Inflammatory Bowel Disease. Gastroenterology, 2022. 162(3): p. 859–876.

7. Ashton, J.J., et al., Genetic Sequencing of Pediatric Patients Identifies Mutations in Monogenic Inflammatory Bowel Disease Genes that Translate to Distinct Clinical Phenotypes. Clinical and Translational Gastroenterology, 2020. 11.

8. Zhou, D., et al., A phenome-wide scan reveals convergence of common and rare variant associations. Genome Medicine, 2023. 15(1).

9. Dickson, S.P., et al., Rare Variants Create Synthetic Genome-Wide Associations. Plos Biology, 2010. 8(1).

10. Goldstein, D.B., The Importance of Synthetic Associations Will Only Be Resolved Empirically. Plos Biology, 2011. 9(1).

11. Wray, N.R., S.M. Purcell, and P.M. Visscher, Synthetic Associations Created by Rare Variants Do Not Explain Most GWAS Results. Plos Biology, 2011. 9(1).

12. Bail, P., How Life Works:A User’s Guide to the New Biology. 2023.

13. Noble, D., It’s time to admit that genes are not the blueprint for life. Nature, 2024. 626: p. 254–255.

14. Vergara-Lope, A., et al., Linkage disequilibrium maps for European and African populations constructed from whole genome sequence data. Sci Data, 2019. 6(1): p. 208.

15. Zhang, W.H., et al., Properties of linkage disequilibrium (LD) maps. Proceedings of the National Academy of Sciences of the United States of America, 2002. 99(26): p. 17004–17007.

16. Stafford, I.S., et al., Supervised Machine Learning Classifies Inflammatory Bowel Disease Patients by Subtype Using Whole Exome Sequencing Data. J Crohns Colitis, 2023. 17(10): p. 1672–1680.

17. Seaby, E.G., et al., A gene pathogenicity tool ’GenePy’ identifies missed biallelic diagnoses in the 100,000 Genomes Project. Genet Med, 2024: p. 101073.

18. Bycroft, C., et al., The UK Biobank resource with deep phenotyping and genomic data. Nature, 2018. 562(7726): p. 203-+.

19. Szustakowski, J.D., et al., Advancing human genetics research and drug discovery through exome sequencing of the UK Biobank. Nature Genetics, 2021. 53(7): p. 942–948.

20. Sollis, E., et al., The NHGRI-EBI GWAS Catalog: knowledgebase and deposition resource. Nucleic Acids Research, 2023. 51(D1): p. D977–D985.

21. Consortium, G.T., The GTEx Consortium atlas of genetic regulatory effects across human tissues. Science, 2020. 369(6509): p. 1318–1330.

22. Rentzsch, P., et al., CADD: predicting the deleteriousness of variants throughout the human genome. Nucleic Acids Research, 2019. 47(D1): p. D886–D894.

23. Horowitz, J.E., et al., Mutation spectrum of reveals recessive inheritance as a main driver of Early Onset Crohn’s Disease. Scientific Reports, 2021. 11(1).

24. Lai, M.H.C., Bootstrap Confidence Intervals for Multilevel Standardized Effect Size. Multivariate Behavioral Research, 2021. 56(4): p. 558–578.

25. Mann HB, W.D., On a test of whether one of two random variables is stochastically larger than the other. The Annals of Mathematical Statistics, 1947. 18(1): p. 50–60.

26. Fay, M.P. and Y. Malinovsky, Confidence intervals of the Mann-Whitney parameter that are compatible with the Wilcoxon-Mann-Whitney test. Statistics in Medicine, 2018. 37(27): p. 3991–4006.

27. Pedregosa, F., et al., Scikit-learn: Machine Learning in Python. Journal of Machine Learning Research, 2011. 12: p. 2825–2830.

28. Agrawal, R., Imieliński, T., & Swami, A., Mining association rules between sets of items in large databases. ACM SIGMOD Record, 1993. 22(2): p. 207–216.

29. Huang, L.S., et al., A fast algorithm for mining association rules. Journal of Computer Science and Technology, 2000. 15(6): p. 619–624.

30. Hahsler, M., B. Grün, and K. Hornik, *arules -: A computational environment for mining association rules and frequent item sets*. Journal of Statistical Software, 2005. 14(15).

31. Hahsler, M., arulesViz: Interactive Visualization of Association Rules with R. R Journal, 2017. 9(2): p. 163–175.

32. Frankish, A., et al., GENCODE: reference annotation for the human and mouse genomes in 2023. Nucleic Acids Research, 2023. 51(D1): p. D942–D949.

33. Sisu, C., GENCODE Pseudogenes. Pseudogenes, 2 Edition, 2021. 2324: p. 67–82.

34. Zheng, D.Y., et al., Pseudogenes in the ENCODE regions:: Consensus annotation, analysis of transcription, and evolution. Genome Research, 2007. 17(6): p. 839–851.

35. Tanwar, V.S., et al., Palmitic Acid-Induced Long Noncoding RNA Regulates Inflammation via Interaction With RNA-Binding Protein ELAVL1 in Monocytes and Macrophages. Arteriosclerosis Thrombosis and Vascular Biology, 2023. 43(7): p. 1157–1175.

36. Honjo, H., et al., RIPK2 as a New Therapeutic Target in Inflammatory Bowel Diseases. Frontiers in Pharmacology, 2021. 12.

37. de Lange, K.M., et al., Genome-wide association study implicates immune activation of multiple integrin genes in inflammatory bowel disease. Nature Genetics, 2017. 49(2): p. 256–261.

38. Sitinjak, B.D.P., et al., The Potential of Single Nucleotide Polymorphisms (SNPs) as Biomarkers and Their Association with the Increased Risk of Coronary Heart Disease: A Systematic Review. Vascular Health and Risk Management, 2023. 19: p. 289–301.

39. Lewis, C.M. and E. Vassos, Polygenic risk scores: from research tools to clinical instruments. Genome Medicine, 2020. 12(1).

40. Barrie, W., et al., Ancient DNA reveals evolutionary origins of autoimmune diseases. Nat Rev Immunol, 2024. 24(2): p. 85–86.

