## supplementary tables and materials for "Moving from GWAS signals to rare functional variation in inflammatory bowel disease through application of GenePy2 as a potential DNA biomarker": SUPPLEMENTARY FIGURES.docx

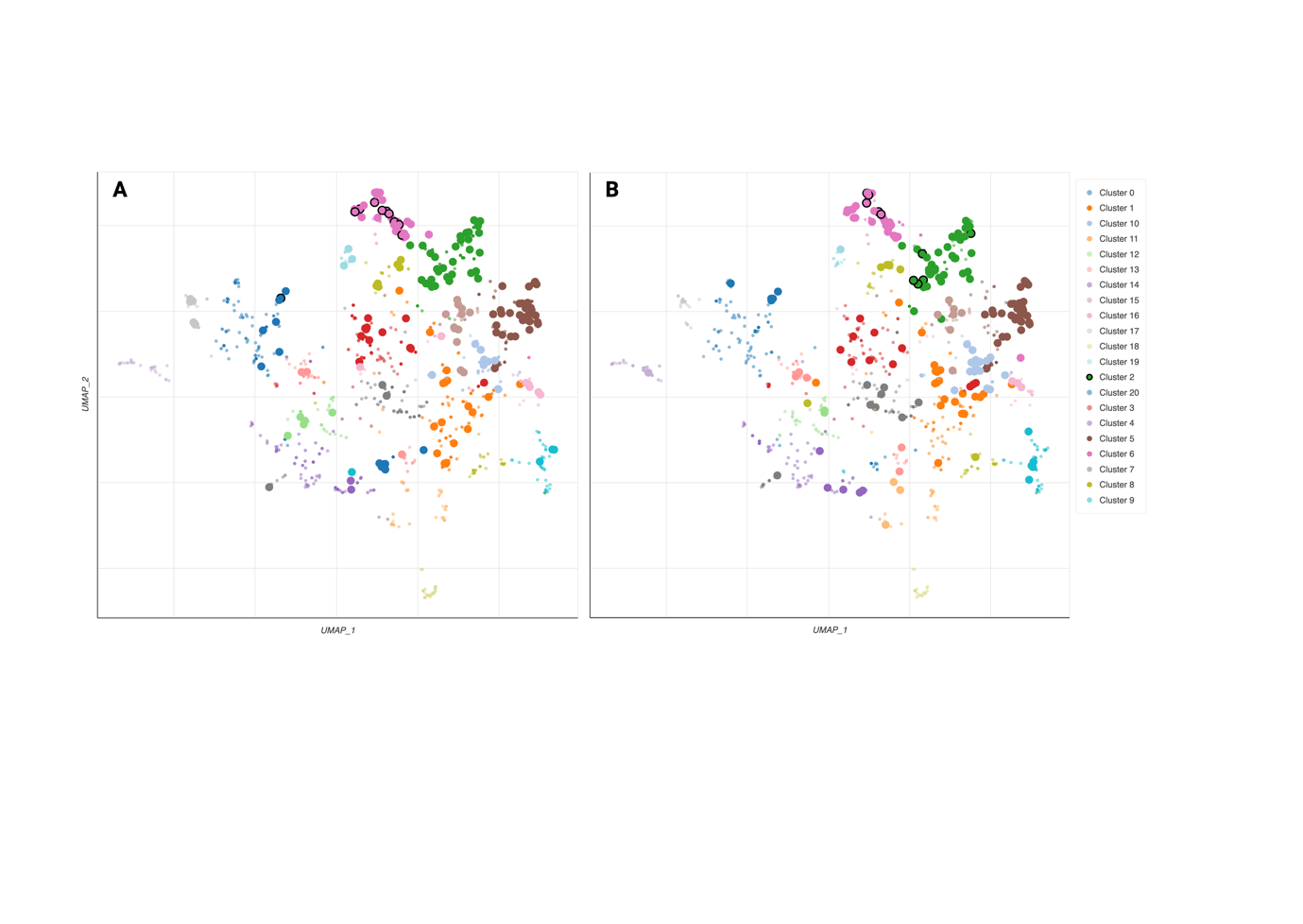

**Figure S1.** Scatterplot of all terms in the WikiPathway_2023_Human gene set library. Each point represents a term in the library. Term frequency-inverse document frequency (TF-IDF) values were computed for the gene set corresponding to each term, and UMAP was applied to the resulting values. The terms are plotted based on the first two UMAP dimensions. Generally, terms with more similar gene sets are positioned closer together. Terms are colored by automatically identified clusters computed with the Leiden algorithm applied to the TF-IDF values. The darker and larger the point, the more significantly enriched the term, and those with black boundaries are the top10 enriched pathways. A). Enrichment by GWAS genes; B). Enrichemnt by monogenic IBD genes.

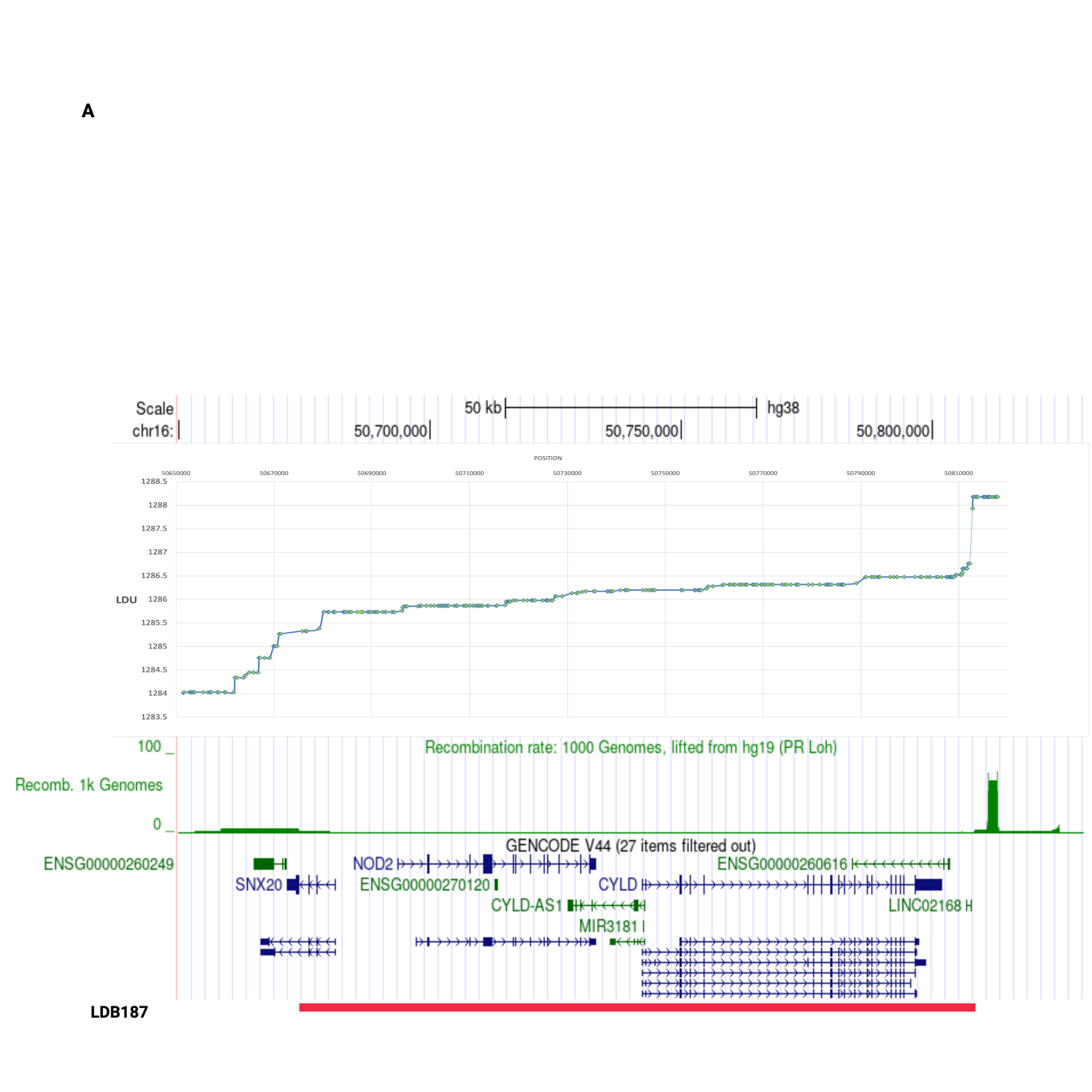

**Figure S2.** The 16q12.1 GWAS association region. Upper panel represents the LDU map of the region with the vertical axis representing the LDU map and horizontal axis representing the physical genomic location. Lower panel represents the recombination rate in the region, followed by the genomic content of the region, including LDB187.

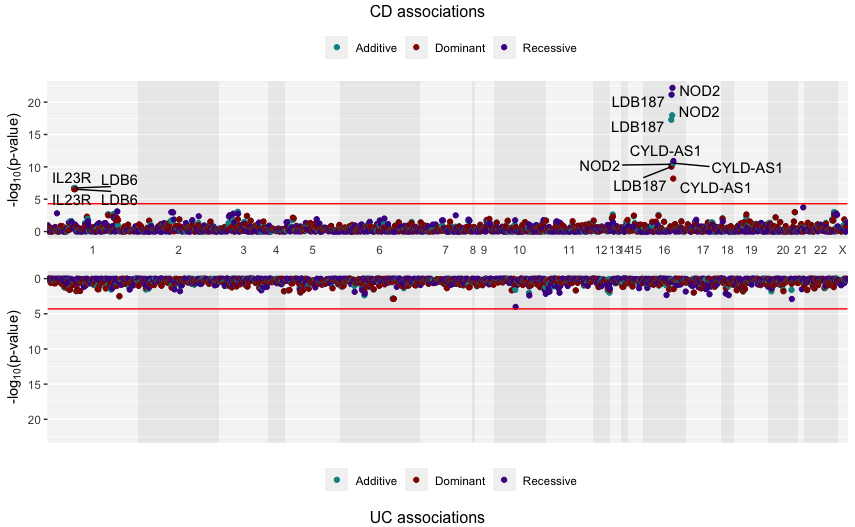

**Figure S3.** Gene Manhattan plots of the association test (Firth’s regression) on gene/LBD’s threshold effects. Individuals were categorised based on the number of pathogenic mutations they have per gene/LDB, adopting different codes for the additive (0/1/2), recessive (0/0/1), and dominant (0/1/1) models for those with 0, 1 and ≥2 mutations.

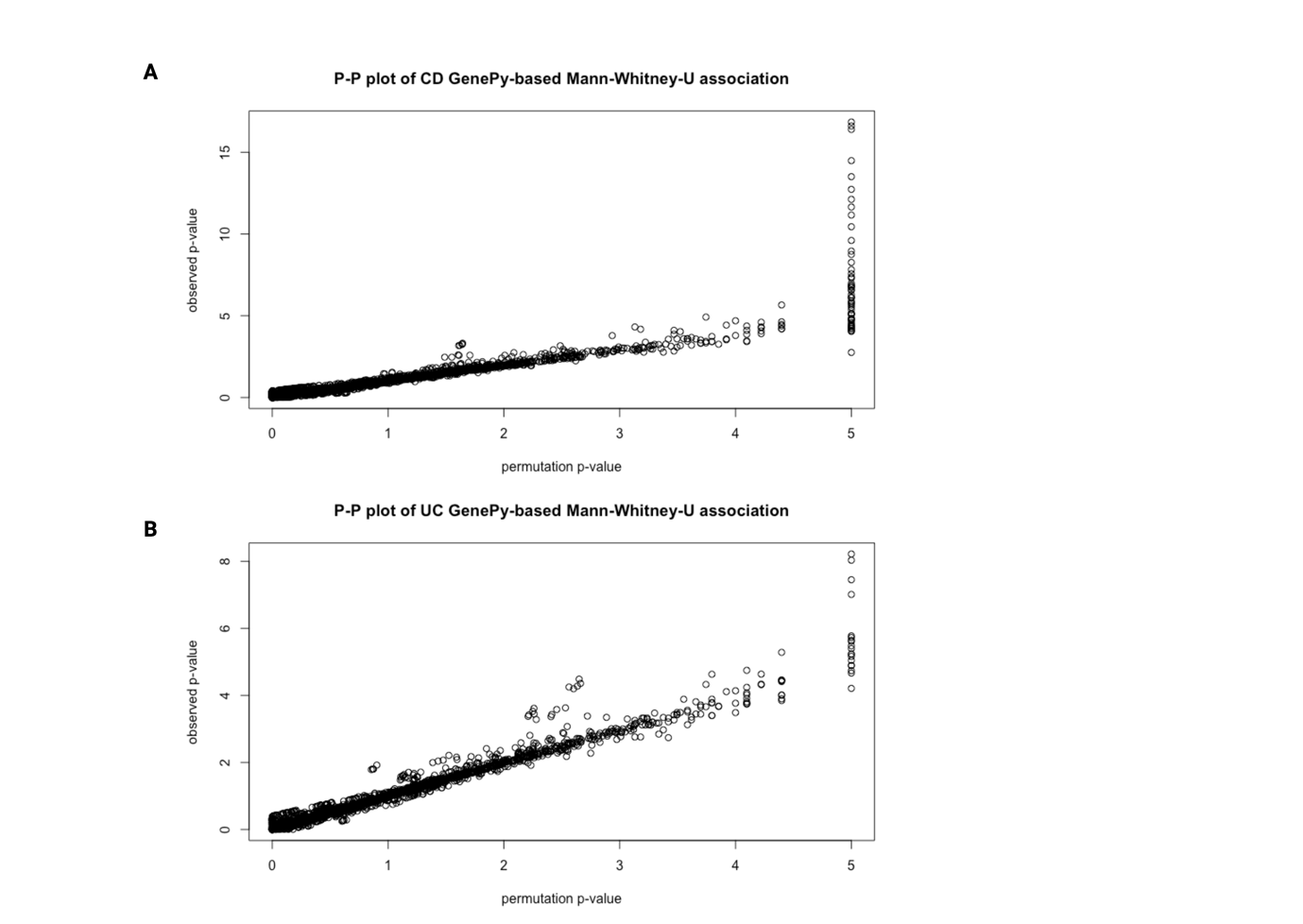

**Figure S4.** P-P plot of GenePy-based associations. X-axis represents permutated p value (n-permutation=10^5^) and Y-axis represents observed p value.
