## supplementary tables and materials for "Moving from GWAS signals to rare functional variation in inflammatory bowel disease through application of GenePy2 as a potential DNA biomarker": Supplementary methods.docx

**Quality control and curation of the IBD cohort based on the UK Biobank data**

On the Exome sequencing data, we filtered for on-target variants with a missingness ratio of less than 10%, prior to genotype-based filtration excluding those with read depth ≤8 and allelic balance ratio <0.15.

Further curation excluded individuals with mismatch self-reported sex versus genetic-sex, >5% discordant exome versus array genotypes, and excessive heterozygote/missingness ratios, adhering to the UK Biobank processing protocol[1]. Participants diagnosed with IBD, UC, or CD were identified using the ICD-10 code. In cases where both CD and UC diagnoses were present, the patient was considered as a CD patient. Individuals without both an immune-related and digestive diagnosis were selected as controls. Subsequently, related cases with second-degree or closer relationships were pruned, retaining the patient with the earliest age at diagnosis, while all related controls were filtered out. To simplify the LD mapping process and focus on the major ethnicity group, the analysis was confined to genetically confirmed White Europeans.

**GenePy2 calculation**

The CADD raw score, derived from a machine-learning-based variant effect predictor, is calculated for each alternative allele based on multiple functional evidence extracted from Ensemble Variant Effect Predictor (VEP) v109 [2]. Genomic coordinates for the genes are determined using GENCODE V43[3], with a padding of 50bp on each side of the defined boundary of genes and the coordinates of the LDB follow the previously described method interpolating from the LDU map to the physical map (HG38). All small variants that pass the QC filtration, including insertions/deletions < 50bp, multi-allelic variants with fewer than 10 alternative alleles, and variants suggested to be pathogenic with a CADD_phred_score_ ≥15, within the gene or LDB were integrated into the GenePy2.0 score and other burden-based tests.

**Burden and threshold tests**

Pathogenic variants (CADD_Phred_Score_≥15) in candidate genes/regions were subjected to a a threshold- and burden-based tests. Firstly, individuals were categorised based on the number of pathogenic mutations they have per gene/LDB, adopting different codes for the additive (0/1/2), recessive (0/0/1), and dominant (0/1/1) models for those with 0, 1 and ≥2 mutations. Subsequently, Firth’s logistic regression test followed by 10^5^ permutations with phenotype resampling was applied to assess the impact of mutations[4]. Secondly, the SKAT-O test, an optimized locus-based burden test, was implemented, which evaluates the burden of mutations while incorporating the top 10 principal components (PCs) of the cohort as covariates [5]. The SKAT-O test used RVTESTs[27] and the regression tests were based on R.

### **Reference:**

1.	Szustakowski, J.D., et al., *Advancing human genetics research and drug discovery through exome sequencing of the UK Biobank.* Nature Genetics, 2021. **53**(7): p. 942-948.

2.	McLaren, W., et al., *The Ensembl Variant Effect Predictor.* Genome Biology, 2016. **17**.

3.	Frankish, A., et al., *GENCODE: reference annotation for the human and mouse genomes in 2023.* Nucleic Acids Research, 2023. **51**(D1): p. D942-D949.

4.	Puhr, R., et al., *Firth's logistic regression with rare events: accurate effect estimates and predictions?* Statistics in Medicine, 2017. **36**(14): p. 2302-2317.

5.	Lee, S., et al., *Optimal Unified Approach for Rare-Variant Association Testing with Application to Small-Sample Case-Control Whole-Exome Sequencing Studies.* American Journal of Human Genetics, 2012. **91**(2): p. 224-237.
